# Right Versus Left Ventricular Myofiber Orientation in Various Congenital Heart Diseases Analyzed with X-Ray Phase-Contrast Tomography

**DOI:** 10.64898/2026.10.01.26364061

**Authors:** Shunsuke Matsushima, Masato Hoshino, Gen Shinohara, Shuichi Shiraishi, Yasutaka Hirata, Takuro Tsukube, Yoshihiro Oshima, Hironori Matsuhisa

**Affiliations:** Department of Cardiovascular Surgery, Kobe Children’s Hospital, Kobe, Japan; Japan Synchrotron Radiation Research Institute, Hyogo, Japan; Department of Cardiovascular Surgery, Jikei University School of Medicine, Tokyo, Japan; Department of Cardiovascular Surgery, JCHO Kyushu Hospital, Kitakyushu, Japan; Division of Thoracic and Cardiovascular Surgery, Niigata University Graduate School of Medical and Dental Sciences, Niigata, Japan; Department of Cardiovascular Surgery, National Center of Child Health and Development, Tokyo, Japan; Division of Cardiovascular Surgery, Japanese Red Cross Kobe Hospital, Kobe, Japan

## Abstract

**BACKGROUND:** While the systemic right and left ventricles have different functional prognoses, only a limited number of reports have addressed their myocardial architecture. This study analyzed myofiber orientation of human specimens with various congenital heart diseases using X-ray phase-contrast tomography.

**METHODS:** Three specimens of each normal heart, ventricular septal defect, right atrial isomerism, left atrial isomerism, and ventricular inversion were examined. In reconstructed images of both ventricles, the center of the free wall was resliced parallel to the myocardial surface. A myofiber helical angle was identified in each slice and plotted from epicardium to endocardium. The plotted curves were compared between both ventricles.

**RESULTS:** In all ventricles, the helical angle was changed from left-handed on epicardial slices to right-handed on endocardial slices. Whereas the right ventricle’s helical angle curves showed a plateau with or without a rapid rise on the endocardial side, the left ventricle’s curves had sustained increase with or without a plateau in the middle. The median wall-thickness– scaled helical angle change rates were 53° and 102° in the right and left ventricle, respectively, and their distribution of each specimen was higher in the left ventricle than in the right ventricle. **CONCLUSIONS:** Ventricular myofiber orientation has a consistent transmural helical transition independently of cardiac morphology and looping. Their transitions can have a plateau and rapid changes permitting the identification of layers within the ventricular wall. The left ventricle has more helical fibers and the right ventricle has more circumferential fibers, which may explain their difference of ventricular performance.

## INTRODUCTION

While long-term survival for complex congenital heart diseases (CHD) has improved over the years, there are an increasing number of patients who need constant specialized care.^1^ The systemic right ventricle (RV) is one of their issues.^2, 3^ The systemic ventricle in patients with transposition of the great arteries following atrial switch repair, congenitally corrected transposition of the great arteries, and single ventricle with RV dominance can fail to sustain the systemic circulation, and there is increasing evidence supporting poor long-term prognosis for each of them.^2, 3^ Their progressive ventricular dysfunction is multifactorial and seems to be associated with intrinsic structural abnormalities as the systemic ventricle.^2, 4^ The myocardial architecture and atrioventricular valve of the RV may be adapted differently to pressure or volume overload in comparison with the left ventricle (LV).^2–4^

As the myocardium is a highly complex three-dimensional (3D) syncytium, the architectural arrangement of the ventricular myocardium is often expressed as myofiber for structural assessments, and the relationship between myofiber orientation and ventricular performance has been explored by various studies.^5–9^ They have revealed that myocardial helical alignment transitioning transmurally in the ventricular wall results in twist dynamics and its disruption occurs in hearts with cardiomyopathy.^8, 9^ On the same theory, myofiber orientation may also explain different functional prognoses of the systemic RV and LV. However, only a limited number of reports have addressed with myofiber orientation in CHD.^10–12^

Our research group, the SPring-8 Cardiovascular Structure Analyzing Research Group, has examined autopsied human heart specimens with CHD at the microscopic level using X-ray phase-contrast tomography (XPCT) at SPring-8, a third-generation synchrotron radiation facility in Japan.^13–17^ This modality provides 3D high-resolution X-ray images without destroying the properties of the specimen.^18^ While the precise locations of conduction tissues in various CHD mainly have been clarified and reported,^16, 17^ myocardial architecture can also be projected in units of perimysium with the same images created by the XPCT. We measured the direction of perimysium in each image resliced parallel to the myocardial surface and investigated its distribution as myofiber orientation.^6^ In this study, myofiber orientation was compared between the RV and LV in normal hearts and CHD to explore their intrinsic microstructural difference.

## METHODS

### Specimen Selection

Human heart specimens available from the pathologic collection at the participating hospitals (Kobe Children’s Hospital, Jikei University School of Medicine, National Center for Child Health and Development) were used. All specimens already had several cutting lines for pathological gross examinations. This study was approved by the ethical committees of the participating hospitals (Kobe Children’s Hospital, 26-16, 11 August 2014; Jikei University School of Medicine, 26-163(7668), 6 October 2014; National Center for Child Health and Development, 815, 16 October 2015), thus waiving the requirement of consent from the family of the deceased. For this study, three specimens of each normal heart, ventricular septal defect, right atrial isomerism, left atrial isomerism, and ventricular inversion were examined (Table 1).^14, 16^ Specimens with a rudimentary ventricle on either side were excluded from the selection. In specimens with right or left atrial isomerism, only specimens with right-hand topology were selected for consistency.

**Table 1.** Demographics of the Specimens.

| Specimen | Diagnosis | Year at Autopsy | Age Group | Sex | BW | Other Malformation |
| --- | --- | --- | --- | --- | --- | --- |
| A1 | Normal heart | 2014 | Neonate | Male | 1.9 kg | Small bowel atresia |
| A2 | Normal heart | 2014 | Infant | Male | 5.6 kg |  |
| A3 | Normal heart | 2014 | Neonate | Male | 0.9 kg | Pulmonary hypoplasia |
| B1 | VSD | 1974 | Infant | Male | 2.4 kg | PLSVC, 13 trisomy |
| B2 | VSD | 1979 | Infant | Female | 2.3 kg | Coarctation of the aorta |
| B3 | VSD | 1982 | Infant | Female | 2.7 kg | PLSVC, 13 trisomy |
| C1 | Right atrial isomerism | 1979 | Neonate | Male | Unknown | AVSD, Pulmonary atresia, Supra-cardiac TAPVC |
| C2 | Right atrial isomerism | 1982 | Infant | Male | 4.7 kg | AVSD, Right aortic arch, Supra-cardiac TAPVC |
| C3 | Right atrial isomerism | 1984 | Infant | Male | 3.9 kg | AVSD, Pulmonary atresia, Right aortic arch, PLSVC, Infra-cardiac TAPVC |
| D1 | Left atrial isomerism | 1968 | Infant | Male | 4.5 kg | VSD, IVC interruption |
| D2 | Left atrial isomerism | 1980 | Infant | Male | 2.8 kg | AVSD, Biliary atresia |
| D3 | Left atrial isomerism | 1983 | Infant | Male | 6.5 kg | AVSD, Pulmonary atresia, PLSVC |
| E1 | SLL-type ccTGA | 1970 | Infant | Female | 4.0 kg | VSD, Ebstein anomaly, Coarctation of the aorta |
| E2 | SLL-type ccTGA | 1977 | Neonate | Female | 3.5 kg | VSD, Pulmonary atresia |
| E3 | SLL-type single ventricle | 1975 | Neonate | Male | 3.5 kg | Coarctation of the aorta |

AVSD indicates atrioventricular septal defect; BW, body weight; ccTGA, congenitally corrected transposition of the great arteries; IVC, inferior vena cava; PLSVC, persistent left superior vena cava; TAPVC, total anomalous pulmonary venous connection; VSD, ventricular septal defect.

### X-Ray Phase-Contrast Tomography

The whole heart specimens were imaged by the XPCT using an X-ray Talbot grating interferometer at the beamline BL20B2 in the SPring-8 (Figure 1). The imaging system and processing values have been described in detail previously.^14, 15, 18^ The data were processed by a high-throughput system to create 3D high-resolution images with a voxel size of 25 μm (12.5 μm in specimen A1, A2, and A3), target density range from 0.9 to 1.2 g/cm3, and density resolution of 1 mg/cm3 (Figure 1). In the 3D image datasets, the ventricular wall appeared to be composed almost exclusively of streaky structures with a thickness of 100-300 μm, corresponding to perimysium (Figure 1).^19^

**Figure 1.**
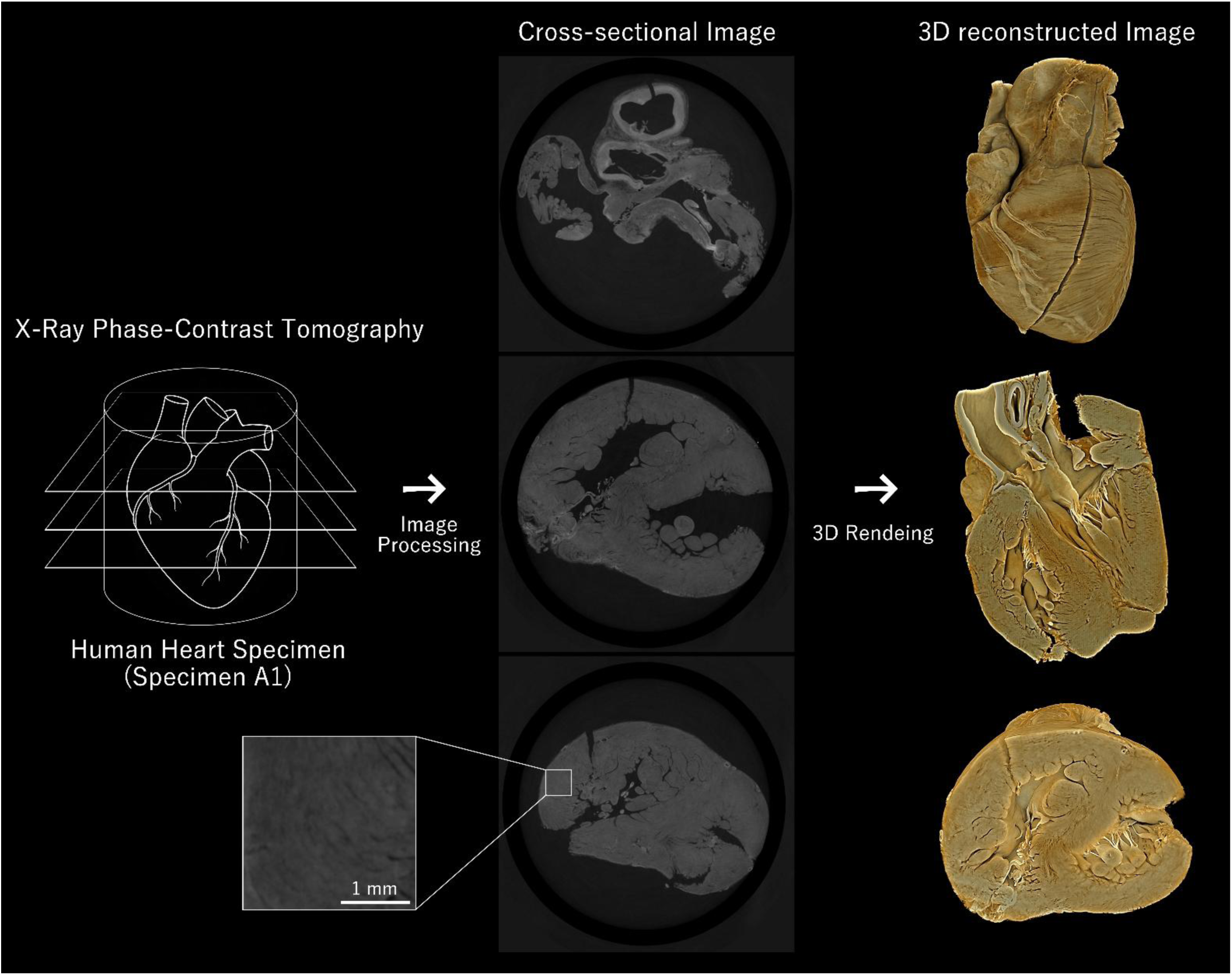
Three-dimensional (3D) Image Processing and Rendering with X-Ray Phase-Contrast Tomography.

### Myofiber Orientation Analysis

Ventricular surfaces were extracted from acquired 3D image datasets and rendered as polygonal meshes by the open-source volume rendering system Drishti (https://github.com/nci/drishti) (Figure 1, 2).^20^ These meshes were cropped and simplified at the center of the free wall on each ventricle, and their normal vectors were derived by the open-source mesh processing system MeshLab (https://www.meshlab.net/) (Figure 2).^21^ The 3D image datasets were resliced with planes orthogonal to the normal vectors, i.e., parallel to the myocardial surface, and trimmed to 5 × 5-mm squares (2.5 × 2.5-mm squares in specimen A1, A2, and A3) by the TransformJ plugin (https://imagescience.org/meijering/software/transformj/) of the open-source image processing software ImageJ/Fiji (https://imagej.net/software/fiji/) (Figure 2).^22, 23^ The number of resliced images of each ventricle was counted and converted to millimeters as a wall thickness. Diameters of streaky structures were measured in the middle slice of these resliced images by the MyofibrilJ plugin (https://imagej.net/plugins/myofibrilj) of ImageJ/Fiji and described with the median value as a myofiber diameter.

**Figure 2.**
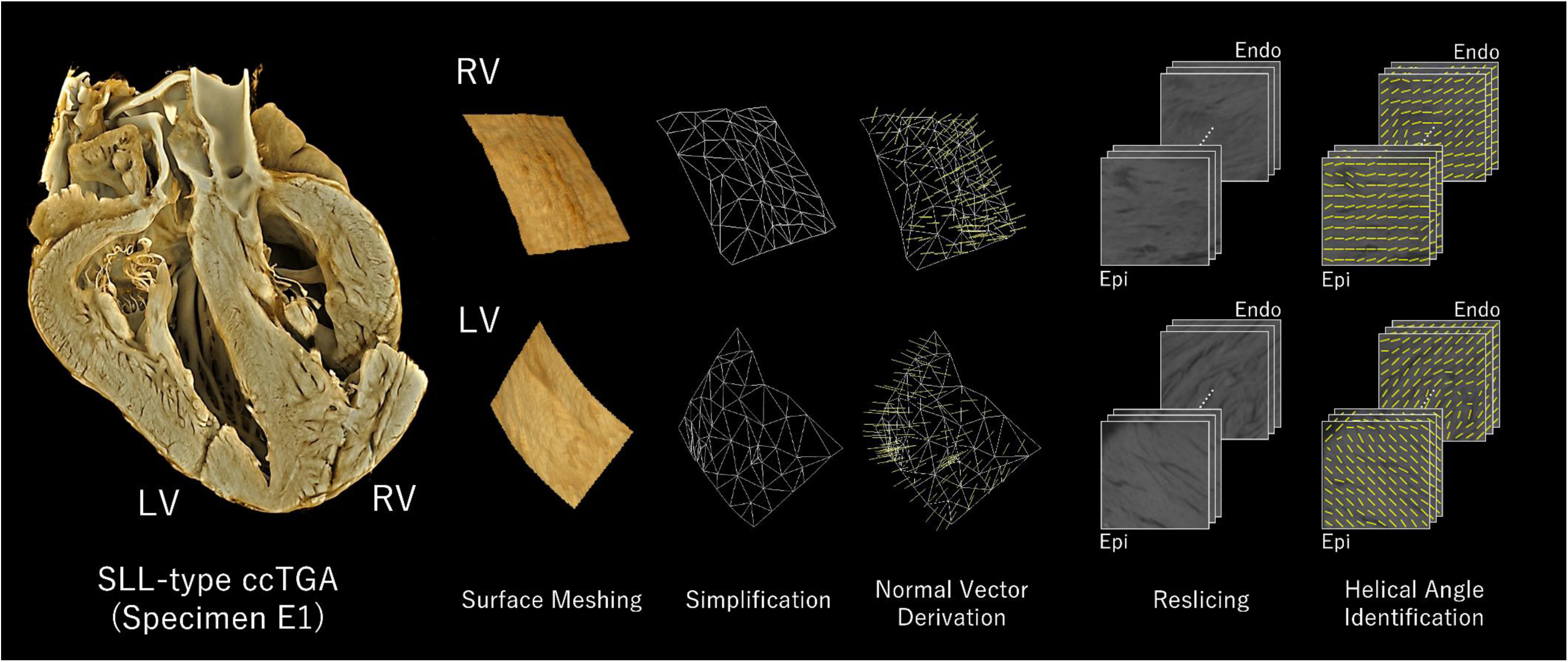
Overview of the Analysis Protocol. After meshing and simplifying the cardiac surface, vectors normal to the center of the ventricular free wall were derived. The image datasets were resliced with planes orthogonal to the normal vectors, and dominant direction was identified as a helical angle in each resliced image. ccTGA indicates congenitally corrected transposition of the great arteries; Endo, endocardial side; Epi, epicardial side; LV, left ventricle; RV, right ventricle.

Dominant direction was identified as a myofiber helical angle in each resliced image by the OrientationJ plugin (https://bigwww.epfl.ch/demo/orientation/) of ImageJ/Fiji (Figure 2).^24^ Because of the difficulty in establishing a common reference axis, these values were adjusted for evaluating relative transmural changes as the mid-range value was set to 0° and right- and left-handed helixes had positive and negative values, respectively. These helical angles were plotted from epicardium (x=0) to endocardium (x=1) and presented with a smooth line curve (Figure 3).^6^ The plotted curves were compared between the RV and LV. Moreover, the range value of the helical angles was presented in each ventricle. The change rates of the helical angles between slices were also calculated in ° per wall thickness and described with the median value and interquartile range.

**Figure 3.**
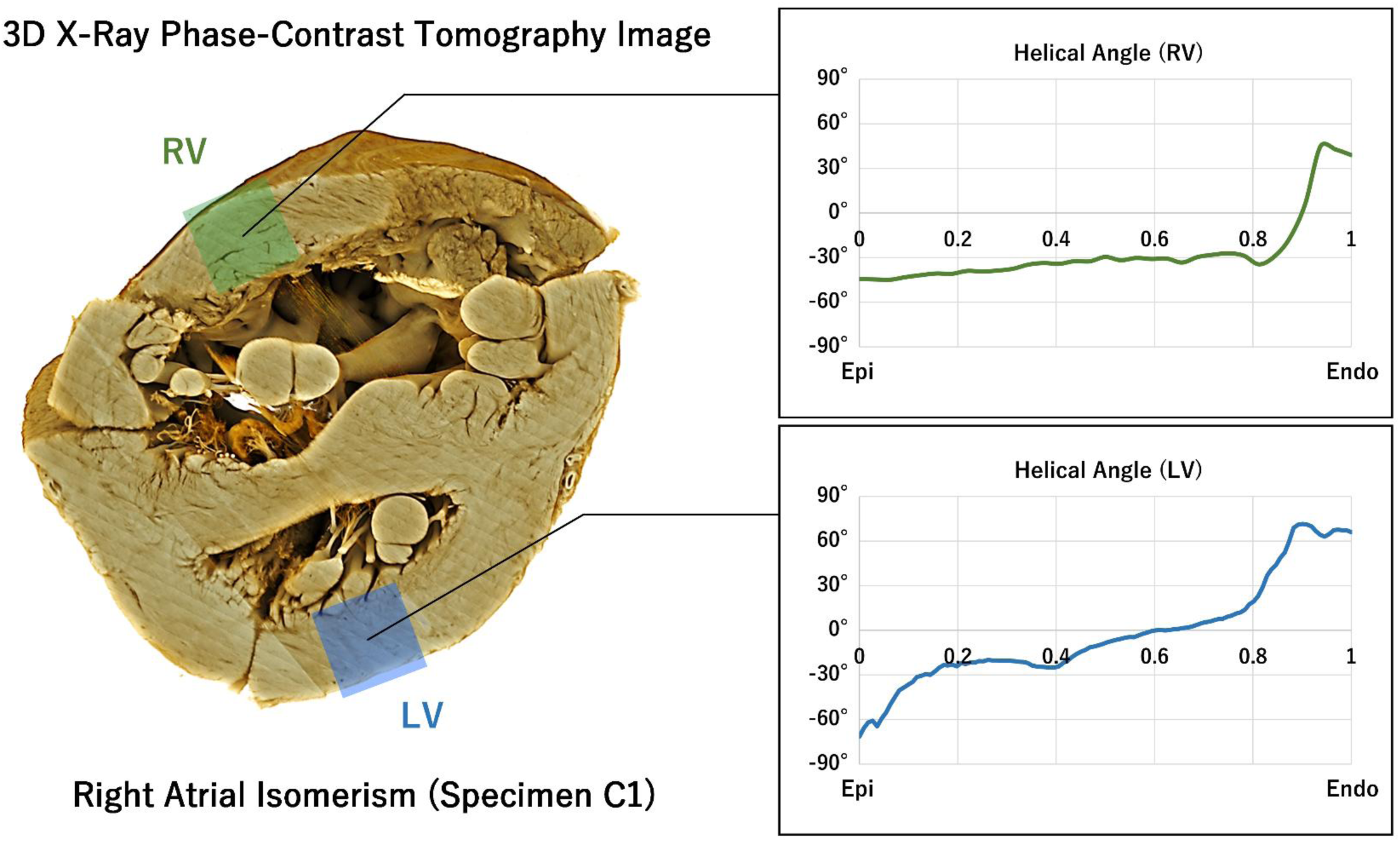
Myofiber Helical Angle Plotting. Helical angles identified in the free wall of each ventricle were plotted from epicardium (x=0) to endocardium (x=1) and presented with a smooth line curve. Endo indicates endocardial side; Epi, epicardial side; LV, left ventricle; RV, right ventricle.

### Statistical Analysis

All statistical calculations were performed with the R environment (version 4.0.3; R Foundation, Vienna, Austria). Continuous variables were compared between the RV and LV in a paired manner for each specimen with the Wilcoxon signed-rank test. The smooth line curve of myofiber helical angles was drawn with Microsoft Office Excel (version 2021; Microsoft Corp., Redmond, WA, USA).

### Data Availability

The data underlying this article will be shared on reasonable request to the corresponding author.

## RESULTS

Wall thickness and myofiber diameter of each analyzed ventricular wall were shown in Table 2. The median value of the wall thickness was 2.3 mm and 4.8 mm in the RV and LV, respectively. The median value of the myofiber diameter was 210 µm and 237 µm in the RV and LV, respectively. The plotted curves of the helical angles were depicted in Figure 4. All ventricles had an identical direction of helical angle change; the helical angle was changed from left-handed on epicardial slices to right-handed on endocardial slices. Whereas the RV helical angle curves showed a plateau with or without a rapid rise on the endocardial side, the LV helical angle curves had sustained increase with or without a plateau in the middle. The range and change rate of myofiber helical angles in each ventricle were shown in Table 2. The median value of the helical angle ranges was 74° and 137° in the RV and LV, respectively. The median value of the wall-thickness–scaled helical angle change rates was 53° and 102° in the RV and LV, respectively, and their distribution of each specimen was higher in the LV than in the RV (Figure 5).

**Figure 4.**
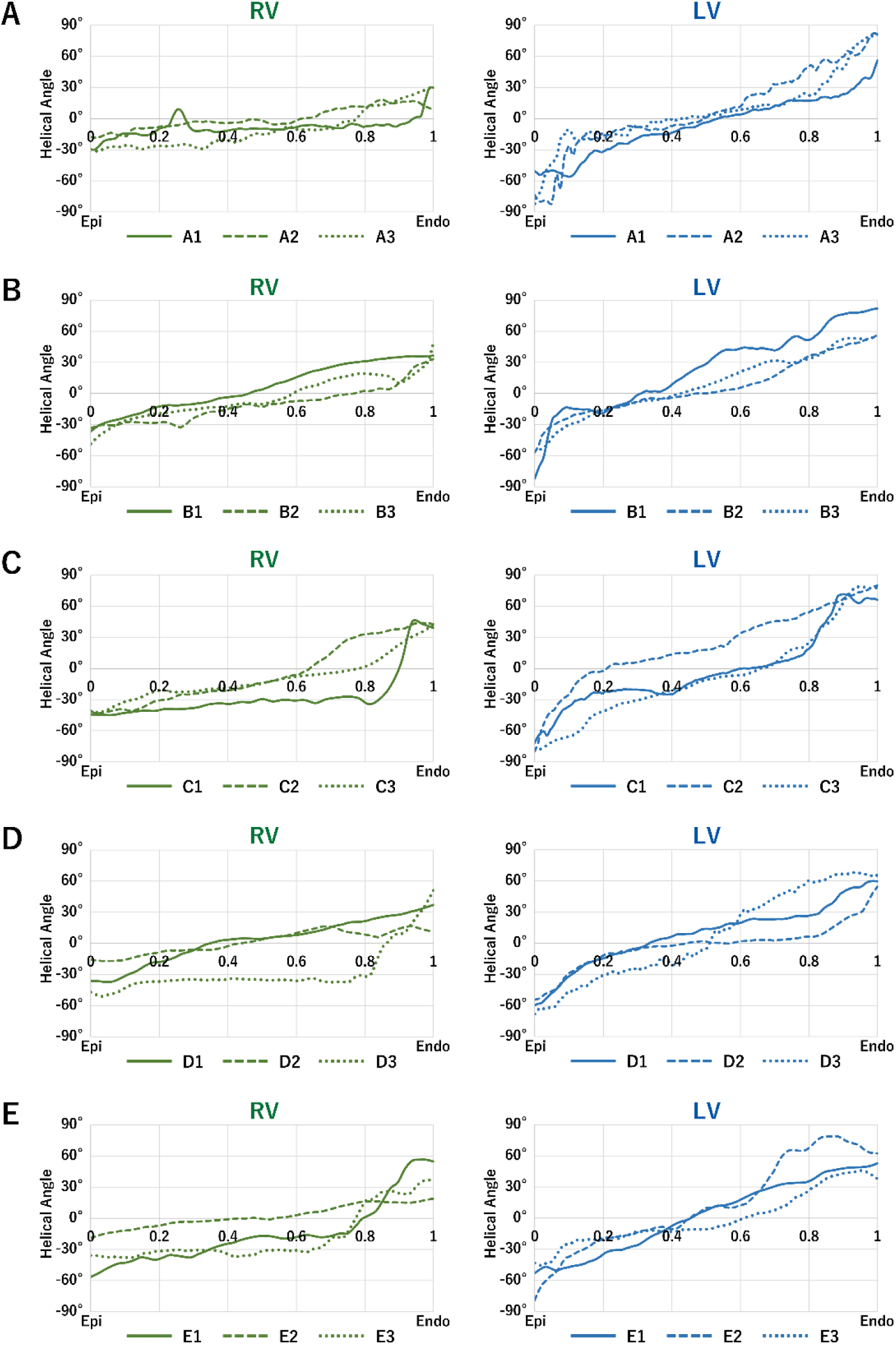
Myofiber Helical Angle Curves. Individual helical angle curves of the RV and LV were plotted for three specimens of each normal heart (A), ventricular septal defect (B), right atrial isomerism (C), left atrial isomerism (D), and ventricular inversion (E). Endo indicates endocardial side; Epi, epicardial side; LV, left ventricle; RV, right ventricle.

**Figure 5.**
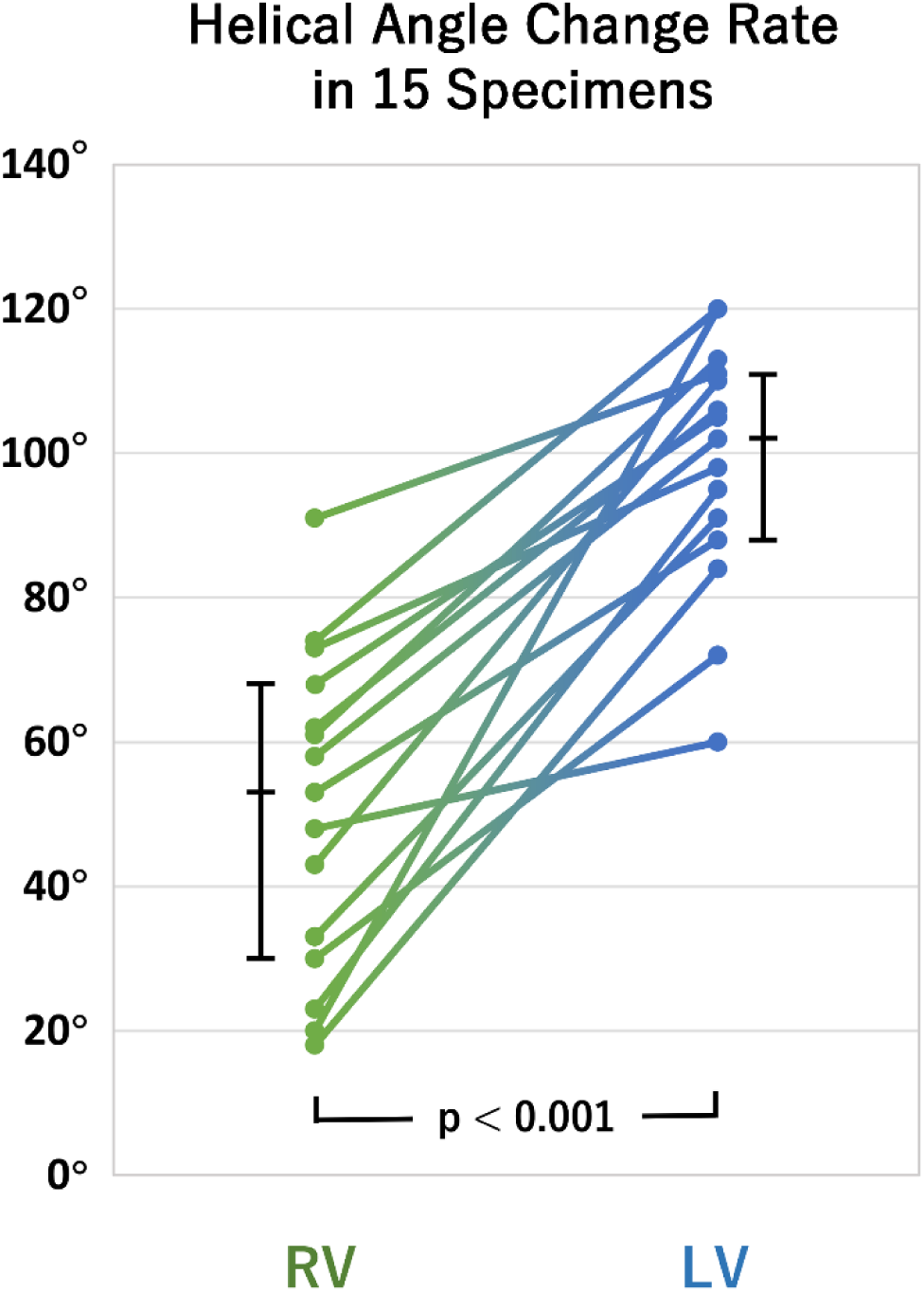
Helical Angle Change Rate Analysis. The change rates of the helical angles between slices were calculated in ° per wall thickness, and their median values were compared between the RV and LV in a paired manner for each of the 15 specimens. LV indicates left ventricle; RV, right ventricle.

**Table 2.** Measured Variables of the Specimens.

| Specimen | Wall thickness, mm | | Myofiber diameter, $\mu\text{m}$ | | Helical angle range, $^{\circ}$ | | Helical angle change rate, $^{\circ}$ | |
| --- | --- | --- | --- | --- | --- | --- | --- | --- |
|  | RV | LV | RV | LV | RV | LV | RV | LV |
| A1 | 2.1 | 3.3 | $112 \pm 44$ | $128 \pm 34$ | 60 | 112 | 18 (-23 - 95) | 84 (4 - 185) |
| A2 | 1.7 | 4.6 | $141 \pm 83$ | $115 \pm 59$ | 37 | 165 | 23 (-29 - 75) | 95 (-19 - 238) |
| A3 | 1.3 | 4.4 | $128 \pm 63$ | $108 \pm 47$ | 63 | 164 | 58 (-3 - 129) | 102 (-55 - 306) |
| B1 | 2.5 | 3.9 | $281 \pm 194$ | $257 \pm 84$ | 73 | 164 | 74 (37 - 107) | 120 (2 - 245) |
| B2 | 3.5 | 4.8 | $142 \pm 29$ | $237 \pm 86$ | 66 | 113 | 53 (4 - 103) | 88 (45 - 143) |
| B3 | 3.5 | 4.8 | $176 \pm 71$ | $226 \pm 36$ | 97 | 110 | 68 (26 - 125) | 105 (73 - 144) |
| C1 | 1.7 | 5.6 | $212 \pm 94$ | $241 \pm 42$ | 90 | 143 | 33 (-14 - 61) | 91 (-10 - 216) |
| C2 | 3.6 | 5.3 | $210 \pm 110$ | $283 \pm 72$ | 88 | 160 | 62 (38 - 134) | 106 (51 - 213) |
| C3 | 1.5 | 5.1 | $209 \pm 63$ | $222 \pm 95$ | 84 | 158 | 61 (33 - 150) | 113 (65 - 200) |
| D1 | 2.3 | 5.7 | $284 \pm 156$ | $232 \pm 89$ | 74 | 120 | 73 (33 - 110) | 98 (45 - 173) |
| D2 | 1.6 | 3.6 | $198 \pm 140$ | $248 \pm 160$ | 34 | 109 | 48 (-14 - 78) | 60 (14 - 169) |
| D3 | 3.1 | 7.6 | $296 \pm 172$ | $282 \pm 153$ | 102 | 137 | 20 (-15 - 123) | 120 (-2 - 254) |
| E1 | 2.5 | 3.2 | $283 \pm 115$ | $231 \pm 99$ | 113 | 106 | 91 (-4 - 149) | 111 (60 - 162) |
| E2 | 2.3 | 4.4 | $290 \pm 90$ | $258 \pm 100$ | 38 | 158 | 43 (16 - 73) | 110 (14 - 250) |
| E3 | 3.6 | 5.3 | $263 \pm 136$ | $272 \pm 109$ | 76 | 92 | 30 (-15 - 106) | 72 (10 - 129) |
| p value | < 0.001 |  | 0.798 |  | < 0.001 |  | < 0.001 |  |

The values of myofiber diameter are mean ± SD. The values of helical angle change rate are expressed in $^{\circ}$ per wall thickness with median (interquartile range). LV indicates left ventricle; RV, right ventricle.

## DISCUSSION

This study explored the arrangement of perimysium as myofiber orientation in a comprehensive series of human heart specimens, encompassing a wide range of cardiac morphology and looping, using the 3D imaging modality with high spatial resolution. It enables the assessment of specimens with thin RV walls, including neonatal ventricles, and allows for sensitive detection of subtle changes in myofiber helical angles. Although these helical angles represent virtual angles measured on a user-defined cross-sectional plane through the complex syncytium, their detailed description can clarify the previously fragmented body of knowledge about myofiber orientation and can provide insights into its previously uncertain clinical significance.^6, 25^

First, the slope of helical angle curves was identical even in hearts with heterotaxy syndrome and ventricular inversion. In other words. ventricular myofiber orientation showed a consistent transmural pattern across different cardiac morphologies and looping patterns. This is entirely consistent with previous gross anatomical and magnetic resonance imaging studies involving similar CHD.^10, 26^ These findings suggest that myofiber orientation is likely determined through a mechanism distinct from cardiac looping and may be attributable to the intrinsic developmental processes of the cardiomyocytes themselves.^10^ Although this remains speculative, the structural discordance between myofiber orientation and cardiac looping may result in the development of deranged cardiomyocyte arrangement and its impairment on the dynamics of LV function.^27^

Second, the plotted curves of helical angles did not only show myofiber orientation arranged in gradual transmural helical transitions but also revealed their varying degrees of steepness. This trend was observed consistently in normal hearts and CHD. Although the myocardial band model has been challenged and largely rejected, the concept of a two-layered RV and a three-layered LV has long been proposed.^6, 25^ The helical angle curves of the present study provide a potential explanation for these previously reported observations as their plateau and rapid change can permit the identification of layers within the ventricular wall.^6^ Although no standardized method for describing myofiber orientation has yet been established, the helical angle curve can be considered a useful approach.^5–7^

Finally, the LV had steeper transmural transition, indicating a smaller proportion of circumferential fibers, than the RV. This means that the LV consists of more helical fibers than the RV even in complex CHD. Helical alignments within the ventricular muscle wall have been considered important in generating a strong contractile force with ventricular twist.^25, 28^ The greater abundance of helical fibers in the LV reflects its role as the principal systolic ventricle. In contrast, the RV appears to contain a greater proportion of circumferential fibers.^12, 25^ Although wall thickness and inherently smaller cardiomyocyte size also play a role, this structural characteristic contributes to the high compliance of the RV for buffering systemic venous return.^4, 29^ These differences in their myocardial architectures and respective physiological roles are likely to underlie the different functional prognoses of the systemic RV and LV. The findings of this study were established by analyzing intrinsic microstructures in neonates or infants and can provide a histological basis for supporting specific surgical procedures such as anatomical repair in patients with congenitally corrected transposition of the great arteries and LV recruitment in patients with borderline left heart disease.^2, 30^ This context may also be relevant to the development of a systemic ventricle through regenerative therapeutic or engineering approaches.^28^

Nevertheless, individual variants that do not conform to these patterns can exist. The RV of specimens B1, D1, and E1 contained a relatively high proportion of helical fibers and may possess the characteristics necessary to function as a systemic ventricle. In some specimens, irregularities were observed in the helical angle curves of the LV, which may indicate deranged myofiber orientation due to the aforementioned mechanism and lead to contractile dysfunction.^8, 9, 27^ This diversity could be one of the factors explaining the variable clinical outcomes following surgical repair of complex CHD and should perhaps be taken into account when estimating their prognoses.^31^

Several limitations of this study should be acknowledged. The available specimens were very limited and obtaining three specimens for each normal heart and CHD was the upper limit in practice. Detailed pre-mortem hemodynamic data were unavailable, and their potential influence could not be accounted for in the analysis. Long-term remodeling effects were also not taken into account because all specimens were from neonates or infants. All specimens had been fixed with formaldehyde and most of them had been stored for a prolonged period, which could have produced differences in terms of the measurements. Due to the pre-existing cutting lines, only the central portion of the ventricular free wall could be analyzed. At a resolution of 25 μm/voxel, blurring of the perimysial boundaries may have resulted in slightly larger myofiber diameters than those reported previously.^19^ In some specimens, the boundary between the endocardium and the papillary muscles was indistinct and required manual delineation. Furthermore, the helical angle calculated in this study represents a structural tensor defined within the plane of the section and differs from the 3D tensor tracts identified by diffusion tensor magnetic resonance imaging.

### Conclusions

Ventricular myofiber orientation is arranged in gradual transmural helical transition with the same direction independently of cardiac morphology and looping. Their helical angle curves can have a plateau and rapid change permitting the identification of layers within the ventricular wall. In both normal hearts and CHD, the LV has more helical fibers and the RV has more circumferential fibers for strong contractility and high compliance, respectively, which may explain the different functional prognoses of the systemic RV and LV.

## Acknowledgments

This study was approved by the SPring-8 Proposal Review Committee (2014B1306, 2015B1370, 2016B1338, 2018A1011, 2018B1003, 2021A1141, and 2021B1293).

